# Networks and clusters of immunometabolic biomarkers and depression-associated features in middle-aged and older community-dwelling US adults with and without depression

**DOI:** 10.1101/2025.07.30.25332419

**Authors:** Asma Hallab, The Health and Aging Brain Study (HABS-HD) Study Team

## Abstract

**Introduction:** Therapy-resistant depression is associated with higher levels of systemic inflammation and increased odds of metabolic disorders. It is, therefore, crucial to identify the biomarkers of high-risk individuals and understand the key features of depression-immune-metabolic networks.

**Methods:** The multiethnic ≥ 50-year-old study population is a subset of the Health and Aging Brain Study: Health Disparities (HABS-HD) study. Spearman’s rank correlation network analysis was performed between immunological, metabolic, and subscales of the Geriatric Depression Scale (GDS). Significant correlations were then evaluated using a multivariable linear regression analysis, including testing for non-linearity and clinical cutoffs.

**Results:** Two clusters were formed: the first included the immune-metabolic biomarkers, and the second included the different subscales of GDS. The two clusters were significantly correlated at six edges. IL-6 and HbA1c were significantly correlated with anhedonic and melancholic features. Abdominal circumference and BMI were significantly correlated with anhedonic features. In the subgroup without current depression, IL-6 and Abdominal circumference maintained a significant edge with anhedonic features. The observed correlations remained statistically significant in the confounder-adjusted regression analysis and followed specific patterns.

**Conclusions:** Symptom clustering showed its superiority over relying on dichotomized depression diagnoses for identifying relevant immunometabolic biomarkers. This study is a first step toward understanding the particularities of immunometabolic depression for better risk stratification and to direct personalized preventive and therapeutic strategies in multiethnic aging populations.

## 1. Introduction

Depression ranks among the top mental health disorders worldwide and is associated with high morbidity and mortality rates. (1, 2) In addition to its related high health costs, one-third of patients with major depressive disorder show a lack of response to adequate treatment strategies and remain therapy-resistant. (3) This non-remission predicts further health complications (4) and social adversities, including unemployment (5) and higher suicide risks. (6–8) Obesity and metabolic disorders are prevalent in patients with depression, particularly in severely affected ones and particularly non-responders. (9) While the effect of depression on metabolic disorders has for long been intuitively associated with the lack of a healthy lifestyle (10) and use of antidepressant medication, (11) recent studies have shown that the causal association is bidirectional, (12) and more complex mechanisms are involved. Low-grade inflammation is commonly described in depression and metabolic disorders, (13) reflecting a latent state of chronic distress and its concomitant impact on the brain and peripheral organs, including the adipose tissue. (14) Our previous study showed that pro-inflammatory cytokines and metabolic biomarkers were significantly higher in middle-aged and older adults with depression. (15) These findings enabled the integration of the immunological biomarkers, primarily Interleukin-6 (IL-6), as statistical mediators within the depression-metabolism crosstalk. (15) A few recent clinical trials investigated causal associations by exploring new treatment pathways targeting anti-inflammatory molecules and evaluating their added effect in treatment-resistant patients with low-grade inflammation. (16, 17) Besides these controversial results, current literature is limited to descriptive and associative studies, and there is a need to understand the underlying pathophysiology by identifying specific clinically relevant features in high-risk individuals, enabling personalized therapeutic strategies. The translation from a generalized paradigm englobing depression into a single diagnostic entity to acknowledging within- and between-individual variability and contesting the potential immunometabolic-symptom specificity allowed initial steps toward understanding the importance of clustering patient subgroups and initiating individualized approaches. (17)

Previous studies dedicated to understanding immune-metabolic networks were explicitly restricted to populations with European ancestry. (18) As such, available findings cannot be generalized to multiethnic populations. Ethnic minorities are exposed to higher adversities and consequently increased risks of affective and stress-related disorders, (19) in addition to higher prevalence of cardiometabolic and immune-mediated pathologies. (14) However, these high-risk groups are less represented in clinical studies. (20) Furthermore, most published studies included broad age ranges, predominantly young and middle-aged populations. (18, 21, 22) However, depression-related symptoms and immunometabolism undergo the dynamic influence of aging and immunosenescence, (23) and combined findings in the young and middle-aged populations might not apply to older adults.

Overall, there is a serious lack of studies on older populations with a multiethnic background. To fill this gap, this study aimed to assess the associations between depression-related features and immunometabolic biomarkers in a multiethnic US population of community-dwelling middle-aged and older adults with and without depression.

## 2. Methods

### 2.1. Study population

The study population is a subset of the Health and Aging Brain Study: Health Disparities (HABS-HD) study, initiated in 2017, to assess health outcomes in Mexican Americans compared to White Americans. (20) It was expanded in 2020 by including Black American participants. Community-dwelling middle-aged and older adults 50 years and older were recruited at the Institute of Translational Medicine at the University of North Texas Health Science Center (UNTHSC). The follow-up had a 24-30-month rhythm.

Since ethnic minorities tend to be rarely involved or minimally represented in clinical studies, the current cohort recruited participants in community-based events and was advertised in social media and newspapers. Participants were eligible if they were at least 50 years old, had no prior diagnosis of dementia (except Alzheimer’s disease), had no Type 1 Diabetes Mellitus (T_1_DM), and have no active cancer during the study or the last 12 months. Furthermore, participants with substance use disorders or severe mental health conditions (except depression and anxiety) were not eligible. Chronic conditions that might impact the cognitive status (exp., late stages of chronic kidney disease, chronic heart failure…) or a history of severe traumatic brain injury were considered exclusion criteria. Participants underwent clinical, neuropsychological, biological, and neuroimaging investigations. The investigations related to the medical history were based on interviews performed by certified specialized clinicians, and participants were asked to disclose current diagnoses and medications. Owing to the community-based aspect of the study, no clinical health records were reviewed.

All procedures contributing to this work comply with the ethical standards of the relevant national and institutional committees on human experimentation and the Helsinki Declaration of 1975, as revised in 2013. The local ethics committee at North Texas provided ethical approval for the source study. Written informed consent was signed by all participants included in the cohort. The current research was performed in compliance with the data use agreement and assessed de-identified aggregated data.

### 2.2. Included variables

#### 2.2.1. Medical history

As previously mentioned, the medical history, current medications, and current relevant diagnoses were collected through interviews performed by certified clinicians, including mental health professionals.

Blood pressure was measured twice at rest. The included systolic and diastolic blood pressure values (SBP and DBP, respectively) were calculated as the mean of the first and second systolic and diastolic blood pressure measurements, respectively. Obesity was defined as a body mass index (BMI = Weight (Kg) / Height (m)^2^) ≥ 30.

Cardiovascular disorders, hypertension, dyslipidemia, and T_2_DM diagnostic criteria were detailed in previous publications. (15) In short, diagnoses were assigned by certified clinicians based on disclosed medical history during the interview, current medication, or abnormal clinical and/or biological findings. (20)

The list of medications was screened case by case to identify antidepressant medication (divided into major groups), benzodiazepines (any type), statins (any type), anti-inflammatory drugs (divided into major groups), and anti-diabetes medication (divided into major groups). One case disclosed a non-specific type of antidepressant medication and was therefore not accounted for.

- **Antidepressant medication** was classified into:

- Selective Serotonin Reuptake Inhibitors (SSRIs): Escitalopram, Citalopram, Sertraline, Fluoxetine, Paroxetine.
- Serotonin-Norepinephrine Reuptake Inhibitors (SNRIs): Duloxetine, Venlafaxine, Desvenlafaxine.
- Norepinephrine-Dopamine Reuptake Inhibitors (NDRIs): Bupropion.
- Tricyclic Antidepressants (TCAs): Doxepin, Amitriptyline.
- Serotonin Antagonist and Reuptake Inhibitors (SARIs): Trazodone.
- Noradrenergic and Specific Serotonergic Antidepressant (NaSSA): Mirtazapine.
- **Anti-inflammatory medications** were classified into:

- All non-steroidal anti-inflammatory drugs (NSAIDs).
- All systemic corticoids (unless details on the administration route are missing).
- All disease-modifying anti-rheumatic drugs (DMARDs) (for rheumatological and systemic disorders, and including a few cases with post-transplantation medication).
- **Anti-diabetes medications** were classified into:

- All oral medications.
- All insulin types.

Medications were reported independently of doses, time, and frequency of intake.

#### 2.2.2. Biological biomarkers

Fasting blood was collected and assessed at the same site. The immunological quantifications of cytokines were performed using the Single Molecule Array (SIMOA) technology developed by Quanterix™, which is a highly sensitive method. C-reactive Protein (CRP) concentrations were measured using an electrochemiluminescent immunoassay on the Meso Scale Discovery (MSD)^®^ platform.

The immunological variables were: Interleukin-5 (IL-5) (pg/mL), Interleukin-6 (IL-6) (pg/mL), Interleukin-10 (IL-10) (pg/mL), Tumor-Necrosis Factor (TNF)-alpha (pg/mL), and CRP (mg/mL).

The metabolic variables were: Triglycerides (mg/dL), Total Cholesterol (mg/dL), High-Density-Lipoprotein (HDL) Cholesterol, Low-Density Lipoprotein (LDL) Cholesterol, HbA1c (%), and BMI.

HbA1c (%) has a strong predictive value on the variations of Glucose (mg/dL) levels over the previous weeks and was therefore preferred for the analysis.

#### 2.2.3. Psychological assessment

Participants underwent neuropsychological tests, including the Geriatric Depression Scale (GDS), (24) administered and analyzed by certified Neuropsychologists.

In this study, for a better interpretation of associations, GDS items were clustered into four clinically relevant subscales:

- <u>**Anhedonia/lack of motivation**</u>: GDS 2 + GDS 4 + GDS 12 + GDS 15 + GDS 20 + GDS 21 + GDS 27 + GDS 28.
- <u>**Melancholia/Negative emotions and cognitions**</u>: GDS 1 + GDS 3 + GDS 5 + GDS 7 + GDS 9 + GDS 10 + GDS 16 + GDS 17 + GDS 19 + GDS 22 + GDS 23 + GDS 25.
- <u>**Worry/Irritability**</u>: GDS 6 + GDS 8 + GDS 11 + GDS 13 + GDS 18 + GDS 24.
- <u>**Cognitive concerns**</u>: GDS 14 + GDS 26 + GDS 26 + GDS 29 + GDS 30.

This approach allows for a better comparability of the results with previous publications. A semantic clustering was preferred over Exploratory Factor Analysis (EFA) and Principal Component Analysis (PCA) since these were rather data-driven and did not allow for a clinically meaningful clustering of GDS items.

Both cases with and without depression were eligible for the study. First, to increase the sample size and statistical power of the results. Second, to adjust for the specific effect of depression and medication in comparison to cases without current depression.

Cases were classified into three groups:

- <u>**Current depression with medication**</u>: Participants disclosed having depression during the interview and are under antidepressant medication.
- <u>**Current depression without medication**</u>: GDS total score ≥ 10 points and no concomitant antidepressant medication.
- <u>**No (current) depression**</u>: GDS total score < 10 points, no disclosed depression, with or without antidepressant medication (in a few cases indicated in sleep disorders in this subgroup).

### 2.3. Statistical analyses

#### 2.3.1. Data distribution

The distribution of the variables was visualized using Q-Q plots and plotted histograms overlaid with corresponding density curves. Normality was statistically assessed using the Shapiro-Wilk test. Skewed variables were log-scaled. Nonparametric methods and dichotomization were applied in cases with persistent skewness.

#### 2.3.2. Missing variables

GDS items were reported as binary variables. Individual responses missing completely at random and corresponding to less than 1% of cases were imputed with the null value. GDS subscales were calculated and reported as continuous variables. No imputation was indicated for the correlation analysis, as this was performed pairwise between available values. Values of variables introduced as confounders in the regression analysis were imputed by the null or median, if they were missing completely at random and presented less than 1% of cases.

#### 2.3.3. Group comparison

The Mann-Whitney, Kruskal-Wallis, and Chi-squared tests were used to compare groups. Results of test statistics and *p-*values were reported.

#### 2.3.4. Correlations

Spearman’s rank correlation was performed pairwise between the immunometabolic biomarkers and the four subscales of GDS as continuous variables. To reduce the risk of Type I error, *p*-values were adjusted for multiple testing using the false discovery rate method (FDR). Statistically significant pairwise correlations (*p_FDR_*-value < 0.05) were visualized in a heatmap matrix where positive correlations were highlighted in red-spectrum colors, and negative correlations with blue-spectrum colors. Statistically non-significant *p_FDR_*-values ≥ 0.05 were crossed out.

Correlation-based network analysis was performed and visualized. Spearman’s coefficient |r|> 0.1 and *p-*value< 0.05 were considered a threshold to visualize relevant statistically significant correlations (|r|≤ 0.1 considered a non-significant correlation). Nodes represented the variables. Edges represented the strength of the correlation and were weighted based on the correlation coefficient value |r|. The distance between the nodes was weighted based on the correlation coefficients. However, the plotting was subject to the constraint of avoiding overlapping and is data- and seed-dependent.

#### 2.3.5. Associative regression analysis

GDS subscales were dichotomized and four dependent variables were considered: “Anhedonia/Lack of motivation” (no=0 / yes=1), “Melancholia/Negative emotions and cognitions” (no=0 / yes=1), “Worry/Irritability” (no=0 / yes=1), and “Cognitive concerns” (no=0 / yes=1). The dependent variables were calculated as binary outcomes, where having at least one point in the corresponding subscore was attributed a positive value, and having no points in the corresponding subscore (GDS subscore=0) was attributed the null value. The independent variables were considered from the immunometabolic cluster. The logistic regression analysis evaluated only variables that survived the adjustment for multiple testing and presented relevant nodes in the correlation-based network analysis, with direct edges to the GDS subscales. GDS subscales without direct correlations with the immunometabolic variables were reported as control groups and for the comparability with previous studies. Independent variables had different distributions; IL-6, HbA1c, and Abdominal circumference were introduced in log-scaled forms to the corresponding models, and BMI was kept in raw form.

Three models were then assessed:

- <u>**Model 1**</u>: adjusted for age (years) + sex (female, male) + ethnicity (“Non-Hispanic White”, “Hispanic”, “Black”) + educational level (years) + and cognitive status (“Normal cognition”, “Mild cognitive impairment”, “Dementia”).
- <u>**Model 2**</u>: Model 1 + cardiovascular diseases (binary) + hypertension (binary) + dyslipidemia-related classes (“No dyslipidemia”, “Dyslipidemia without medication”, “Dyslipidemia with medication”) + T_2_DM-related classes (“No diabetes”, “Diabetes without medication”, “Diabetes with medication”) + use of benzodiazepines (binary) + use of anti-inflammatory medication (“NSAIDs”, “Corticoids”, “DMARDs”) + Alcohol consumption (binary) + current Tobacco smoking (binary).
- <u>**Model 3**</u>: Model 2 + depression-related classes (“No (current) depression”, “Current depression without medication”, “Current depression with medication”).

Variance inflation factor (VIF) was calculated in each model, and only variables with factors around one were included. Furthermore, to reduce the risk of multicollinearity:

- For models where a T_2_DM-related predictor (HbA1c) was introduced as an independent variable, T_2_DM was not included as a confounder.
- For models where the obesity-related predictors (BMI and Abdominal circumference) were introduced as independent variables, obesity was not included as a confounder.

Models 3 were considered the model of choice and visualized, as they accounted for the effects of depression and antidepressant medication in the total population.

Models were assessed for non-linearity along three degrees of freedom (df=3, 4, and 5), and were compared to the linear model. The Akaike Information Criterion (AIC) was used to find the best fit.

To facilitate the interpretation of the findings, the analyses were repeated based on clinical cutoffs (HbA1c, Abdominal circumference, and BMI). IL-6 was not included in this analysis since levels were predominantly much lower than clinical cutoffs. The dichotomized odds ratios were based on model 3, and the reference group included those with levels under the corresponding cutoff value of the studied variable.

#### 2.3.6. Sensitivity analysis

To optimize the interpretability of intricate findings and ensure the comparability of the results with published data, the tertile categorization was then applied, and the previous steps were repeated with the first tertile, T1, as reference.

The statistical analysis was performed using RStudio (Version 2024.12.1, Posit^®^, USA).

## 3. Results

### 3.1. Study Data

Ambiguous BMI values (BMI < 15 and BMI > 70) were considered missing in three cases, particularly for the correlation analysis. These and other missing BMI values were then imputed with the median value, allowing an optimal integration of obesity as a confounder in the regression analysis. Similarly, missing cognition-related diagnoses were imputed by the null in nine cases. The cytokines, CRP, HbA1c, serum lipids, and Abdominal circumference had a right-skewed distribution. BMI had a close-to-normal distribution and was kept in its raw form.

### 3.2. Study population

The study included 3,838 participants aged between 50 and 90 years, with a highly represented multiethnic background (White 35%, Hispanic 37%, and Black 28%). Females presented 62% of the cohort.

### 3.3. Group comparison

The detailed GDS subscores are detailed in Supplementary Figure 1.

**Supplementary Figure 1:**
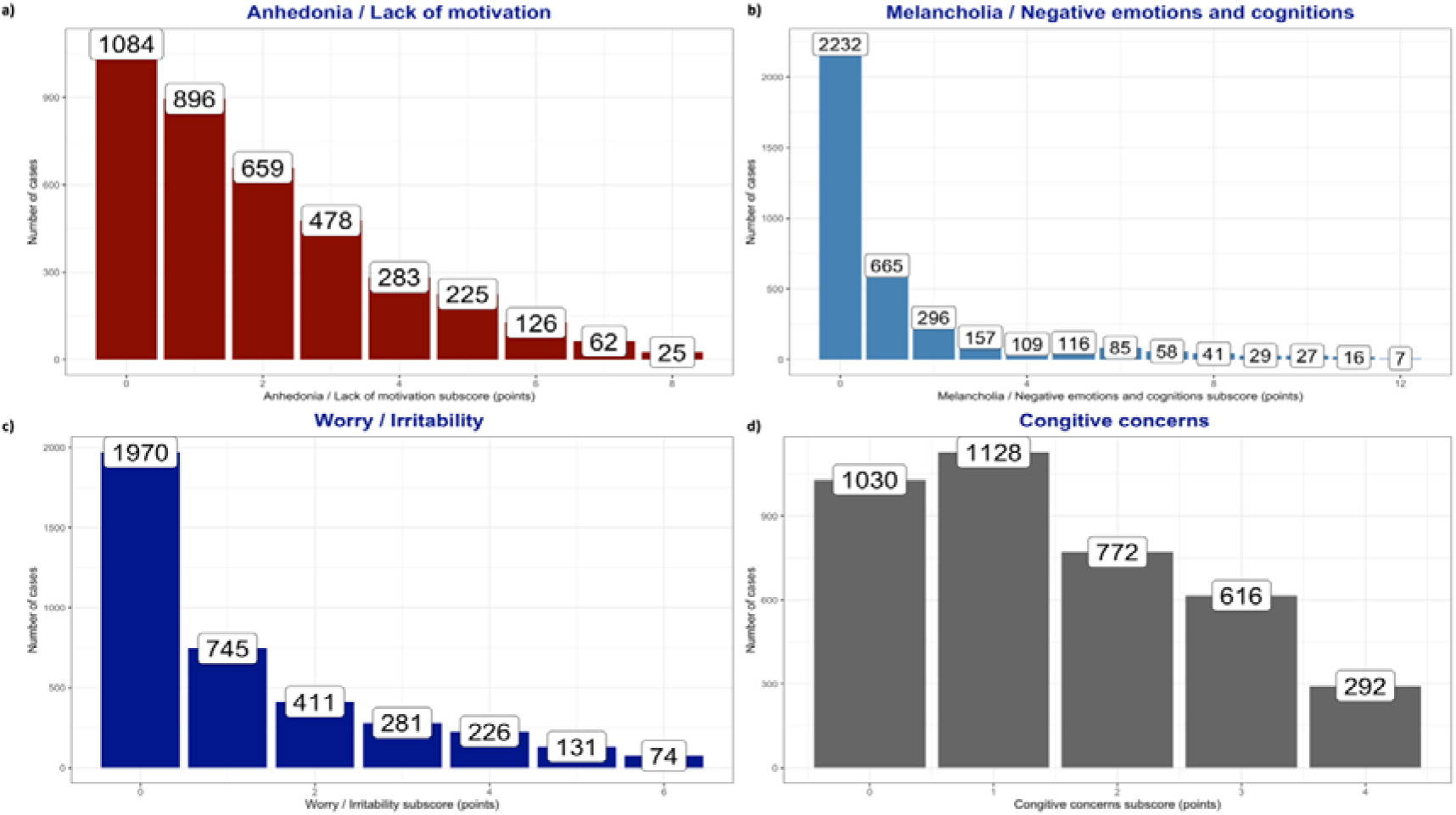
Details of GDS subscores.

Black participants reported more “Anhedonia/Lack of motivation” symptoms, while Hispanic participants reported more “Melancholia/Negative emotions and cognitions”, “Worry/Irritability”, and “Cognitive concerns” (Figure 1.a). Women reported significantly more “Melancholia/Negative emotions and cognitions” and “Cognitive concerns” compared to males (Figure 1.b).

**Figure 1:**
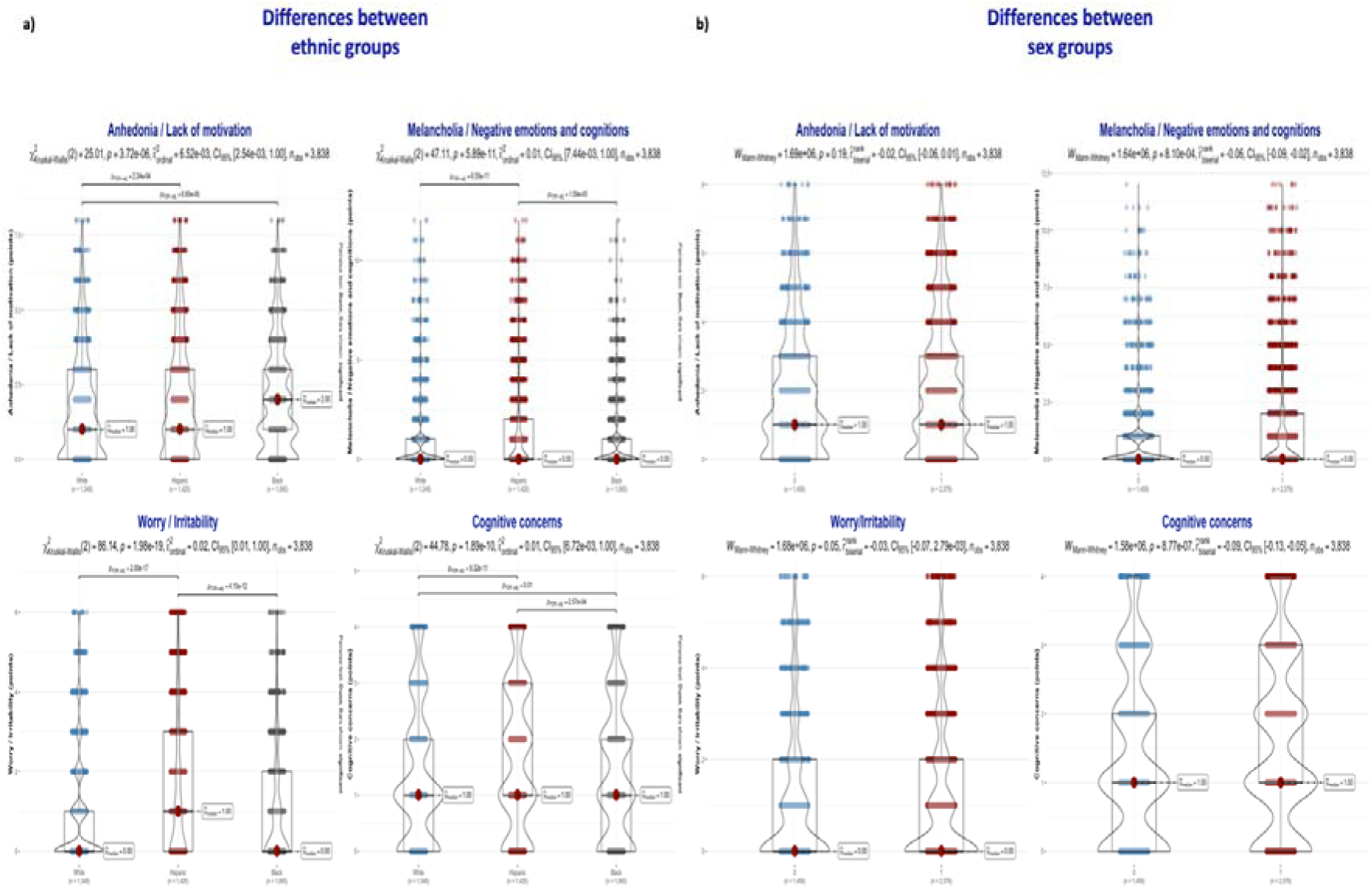
Differences in Geriatric Depression Scale subscores between ethnic and sex groups. **1.a)** Differences between ethnic groups. **1.b)** Differences between sex groups.

Participants with T_2_DM and obesity had significantly higher GDS scores (*p*-values < 0.001). Details on GDS items in T_2_DM and obesity were visualized in Figure 2.

**Figure 2:**
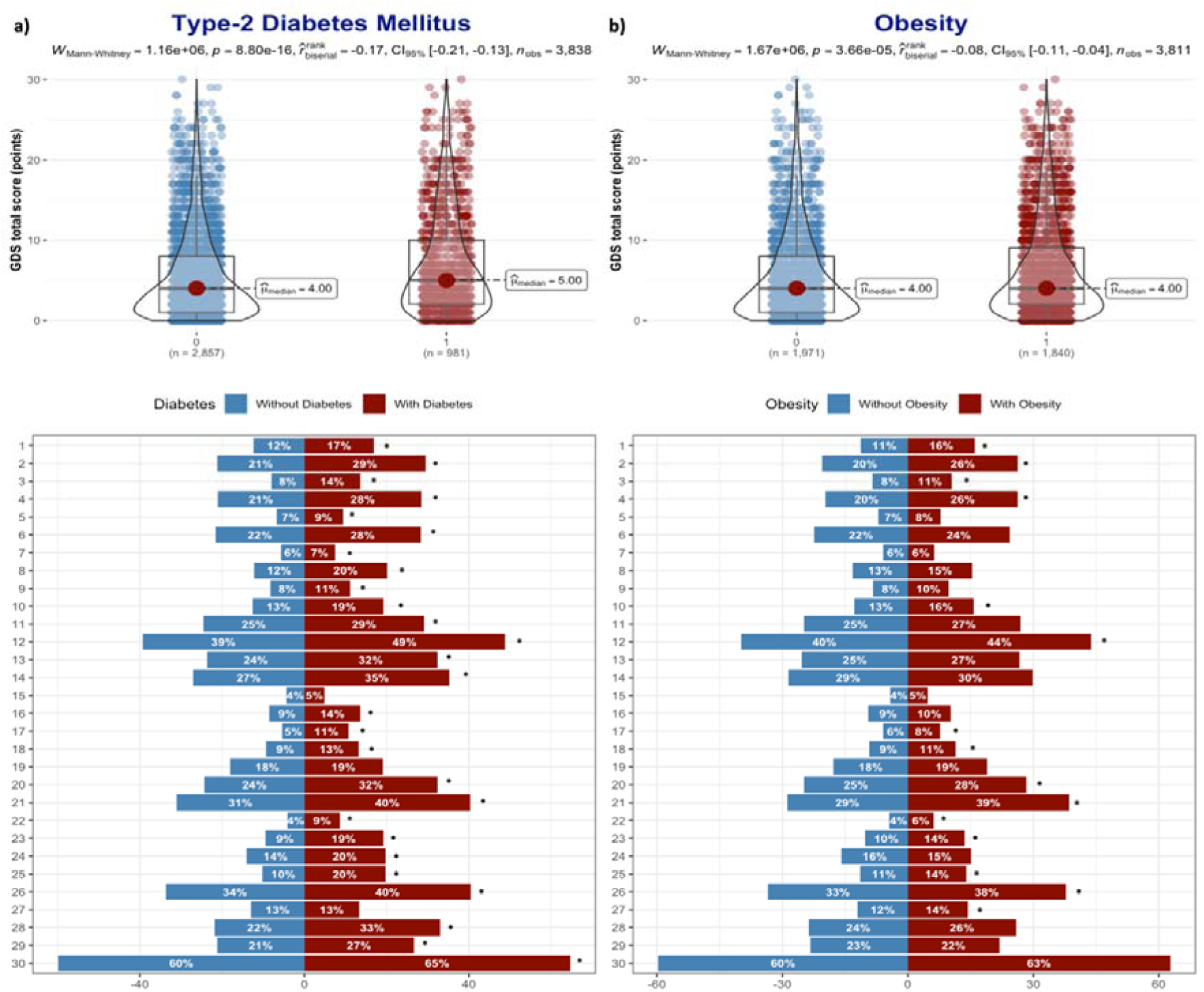
Differences in Geriatric Depression Scale total score and items between participants with and without Type 2 diabetes mellitus and Obesity. **2.a)** Type 2 diabetes mellitus. **2.b)** Obesity. ***Footnote:** * for p-value < 0.05*.

### 3.4. Correlation analysis

A correlation analysis was performed between all relevant variables, and FDR-adjusted results were plotted in a correlation matrix (Figure 3).

**Figure 3:**
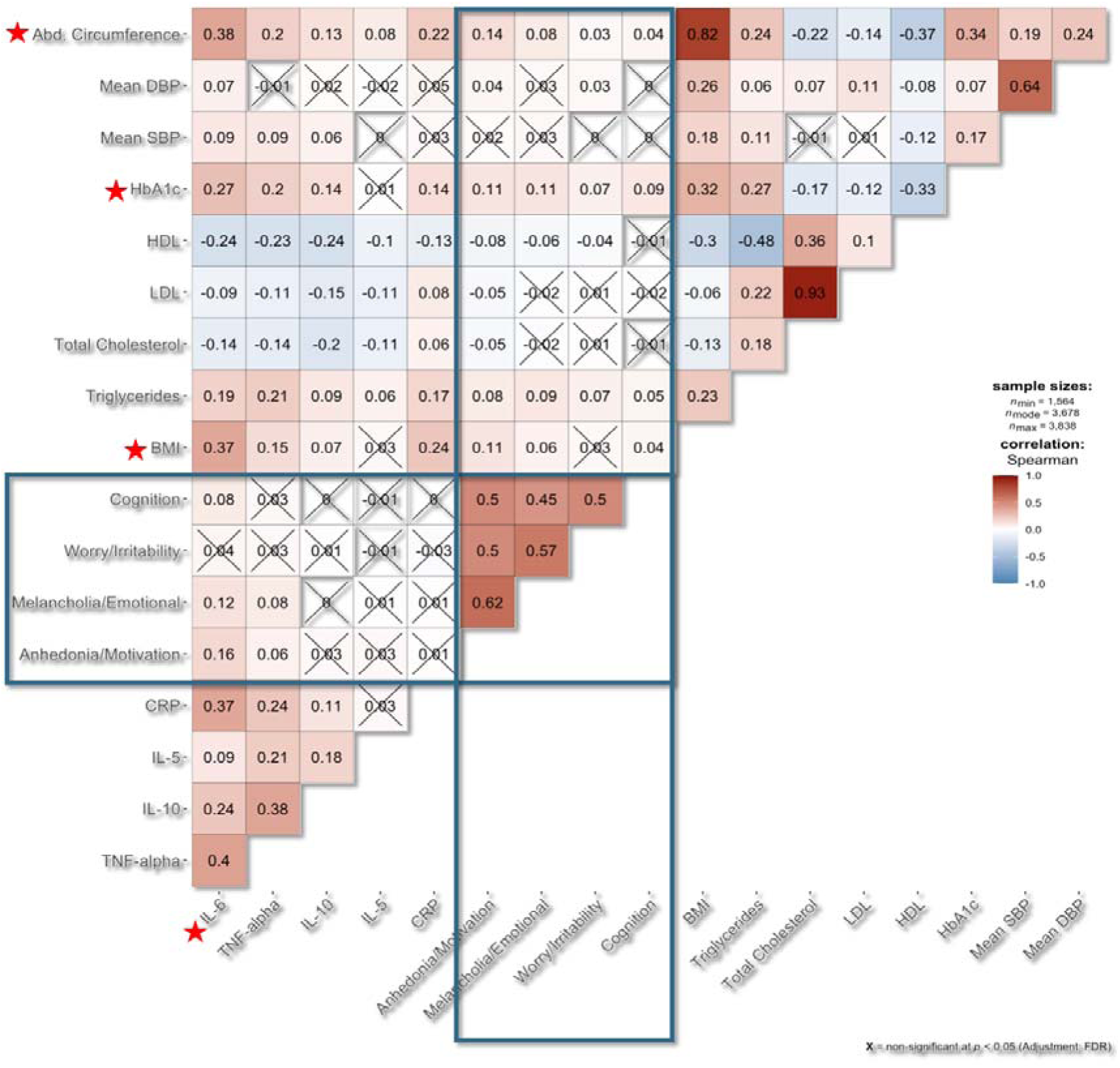
Correlation matrix between immunometabolic biomarkers and GDS subscores with *p*-values adjusted for multiple testing.

In the correlation-based network analysis, two clusters were formed: the first included the immune-metabolic biomarkers, and the second included the different subscales of GDS. The two clusters were significantly correlated at six edges, with four primary nodes of the immunometabolic cluster (IL-6, HbA1c, Abdominal circumference, and BMI) and two nodes of the depression-related features (“Anhedonia/Lack of Motivation” and “Melancholia/Negative emotions and cognition”) (Figure 4.a).

**Figure 4:**
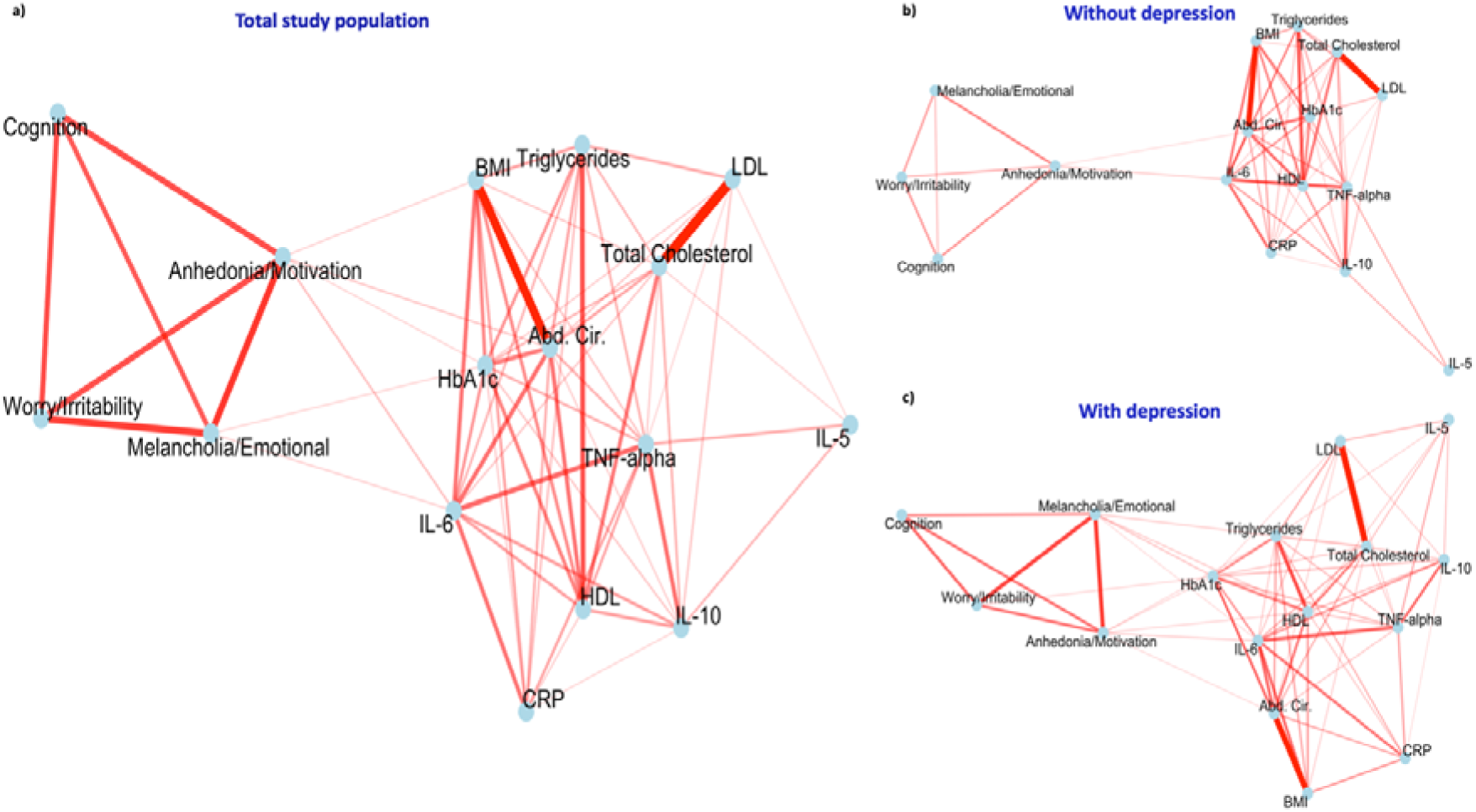
Correlation network analysis in the total study population and stratification by current depression status. **4.a)** Total study population. **4.b)** Without depression. **4.c)** With depression.

Data were stratified by current depression status. Those without current depression maintained two significant edges between IL-6, Abdominal circumference, and “Anhedonia/Lack of motivation” (Figure 4.b). The number of edges between the two clusters increased in the subgroup with current depression and involved additionally lipids (Figure 4.c).

### 3.5. Regression analyses

#### 3.5.1. Assessing linearity in regression models

Different models were tested for non-linearity, and the best fit was kept for the analysis.

Higher IL-6 was significantly associated with higher odds of reporting “Anhedonia/Lack of motivation” and “Melancholia/Negative emotions and cognitions” in a linear trend.

Similarly, higher Abdominal circumference and BMI measures were linearly associated with higher odds of “Anhedonia/Lack of motivation”.

The HbA1c-“Anhedonia/Lack of motivation” and HbA1c-“Melancholia/Negative emotions and cognitions” associations had non-linear patterns with statistical significance at df=4. Similarly, the association between Abdominal circumference and “Cognitive concerns” had a non-linear pattern with statistical significance at df=5.

#### 3.5.2. Independent variables based on clinical cutoffs

HbA1c levels equal to or higher than 6.5% were significantly associated with higher odds of “Anhedonia/Lack of motivation” (OR=1.28 [1.02, 1.60], *p*-value=0.034) and “Melancholia/Negative emotions and cognitions” (OR=1.27 [1.02, 1.57], *p*-value=0.030). Abdominal circumference measures equal to or higher than 40 inches for males /35 inches for females, respectively, were associated with higher odds of “Anhedonia/Lack of motivation” (OR=1.26 [1.06, 1.50], *p*-value=0.009). Similarly, BMI measures equal to or higher than 30 were associated with higher odds of “Anhedonia/Lack of motivation” (OR=1.21 [1.03, 1.42], *p*-value=0.020). The clinical cutoff values of BMI did not explain the association between the variation of BMI and the odds of “Cognitive concerns” at df=5.

#### 3.5.3. Independent variables based on tertiles

This method did not outperform the clinical cutoff-based analysis, particularly for HbA1c, as no significant results were observed.

The association between IL-6 and “Anhedonia/Lack of motivation” confirmed the linear trend as OR in T3 (OR=1.45 [1.12, 1.89], *p*-value=0.005) and T2 (OR=1.40 [1.10, 1.79], *p*-value=0.007) were significantly higher than T1. Only very high values (T3) of Abdominal circumference (OR=1.30 [1.06, 1.59], *p*-value=0.010) and BMI (OR=1.23 [1.01, 1.51], *p*-20 value=0.042) were significantly associated with higher “Anhedonia/Lack of Motivation” compared to the reference.

The association between BMI and “Cognitive concerns” showed that values of BMI in T2 were significantly associated with higher odds of “Cognitive concerns” compared to the reference.

All significant models with plotting of the clinical cutoff-based odds ratio were visualized in Figure 5.

**Figure 5:**
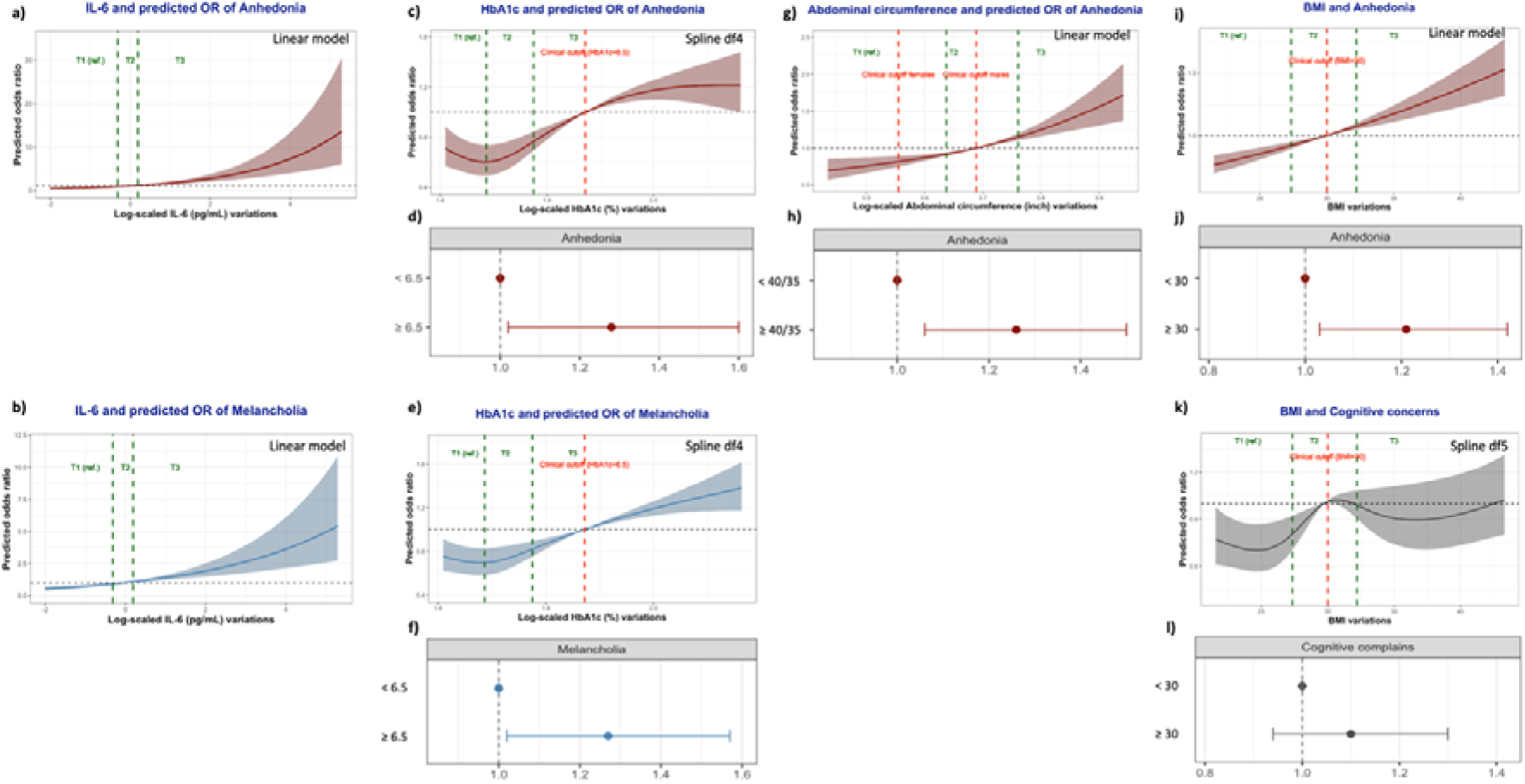
Summary visualization of regression models. **5.a)** IL-6 and predicted odds ratios of anhedonia. **5.b)** IL-6 and predicted odds ratios of melancholia. **5.c)** Glycated hemoglobin A1c and predicted odds ratios of anhedonia. **5.d)** Dichotomized glycated hemoglobin A1c and predicted odds ratios of anhedonia. **5.e)** Glycated hemoglobin A1c and predicted odds ratios of melancholia. **5.f)** Dichotomized glycated hemoglobin A1c and predicted odds ratios of anhedonia. **5.g)** Abdominal circumference and predicted odds ratios of anhedonia. **5.h)** Dichotomized Abdominal circumference and predicted odds ratios of anhedonia. **5.i)** Body mass index and predicted odds ratios of anhedonia. **5.j)** Dichotomized body mass index and predicted odds ratios of anhedonia. **5.k)** Body mass index and predicted odds ratios of cognitive concerns. **5.l)** Dichotomized body mass index and predicted odds ratios of cognitive concerns. ***Footnote:** visualization of x-axis values between 5% and 95%*.

## 4. Discussion

The main outcome of the study was the two correlation-weighted clusters revealed between immunometabolic variables and different GDS subscores, with four relevant points in the immunometabolic cluster (IL-6, HbA1c, Abdominal circumference, and BMI), all of which created significant direct edges with at least one GDS-specific cluster. Only “Anhedonia/Lack of motivation” and “Melancholia/Negative emotions and cognitions” were directly correlated with the immune-metabolic biomarkers in the total population. The stratification by current depression status showed the importance of the correlation between IL-6, Abdominal circumference, and anhedonic features in individuals without current depression. These findings were replicated in the regression analysis after adjusting for multiple confounders. While IL-6 had a linear association with higher odds of anhedonic and melancholic features, clinical cutoffs of other variables showed significant and practically interpretable associations with the same outcomes (“Anhedonia/Lack of motivation” and “Melancholia/Negative emotions and cognitions”). Tertile-based cutoffs allowed the identification of the significant association between medium ranges of BMI and “Cognitive concerns” at df=5, not revealed by the clinical cutoff-based method. However, the intricate associations with “Cognitive concerns” might be confounded by an intermediate effect of the anhedonic or melancholic features, as visualized in the network analysis, and might not represent direct effects.

### 4.1. Clustering depression

Several strategies were evaluated to assess associations and networks between depression-related symptoms, metabolic, and immunological biomarkers. Netherlands Study of Depression and Anxiety (NESDA) included participants with current or past depression diagnoses (42.9±12.9 years, n=2,321) and used the self-rated version of the Inventory of Depressive Symptomatology (IDS-SR). The network analysis provided comparable but also conflicting results to our study. (21) Only IL-6 maintained a direct correlation with the total depression score after adjusting for confounders. In adjusted models where only CRP was evaluated, this was only directly correlated with the “energy level” subscore. Introducing the inflammatory biomarkers in a network with adjusted and regularized models showed no direct association between these and depression subscores. In less conservative adjusted networks without regularization, several inflammation-related edges were statistically significant. The corresponding symptoms were mainly related to sleep, energy, pain, and irritability. (21) Although sleep and pain are not investigated in GDS items, irritability and worry did not show any significant association with the immunometabolic biomarkers in our study. The association between sleep disorders and dyslipidemia has been commonly described in older populations; (25) however, there is still a need to understand how depression-related sleep disorders, inflammation, and dyslipidemia are associated.

In a German cohort of middle-aged patients with T_1_DM and T_2_DM (44.9±13.9 years, n=1,260), several immunological biomarkers were tested for their association with different depression features. (26) Depression symptoms were assessed by the Center for Epidemiological Studies-Depression Scales (CES-D), and the subscores were divided into cognitive-affective, somatic, and anhedonic. The study found significant associations between depression symptoms and some immunological biomarkers; none were included in our study. However, the German study has some limitations, since the exploration of a high panel of immunological biomarkers without adjusting for multiple testing exposes the data to high risks of Type I error. Out of 76 biomarkers, only nine were associated with depression in general, and two to six with each cluster (four with cognitive-affective, six with somatic, and two with anhedonia). The associations were, however, stronger in the T_2_DM than the T_1_DM subgroup after stratification. (26) In our study, we followed a selective approach based on the correlation network and strength of the correlation coefficients, in addition to adjusting for multiple testing. This allowed focusing on variables directly correlated with GDS subscores. While T_1_DM was not assessed in our cohort, cases with T_2_DM differentiated remarkably in their GDS items from those without diabetes and obesity-related grouping.

Older studies tended to classify depression into “melancholic” versus “atypical”. Findings from the Netherlands study on Depression and Anxiety in middle-aged adults (41.3±14.6 years, n=776) suggested that “melancholic depression” was correlated with hypothalamic-pituitary-adrenal axis hyperactivity (higher cortisol levels), while “atypical depression” was rather correlated with inflammatory and metabolic dysregulations. (27) Considering “anhedonia” as the modern label of “atypical depression”, both melancholic and atypical features were associated with the immunometabolic biomarkers in our study, which contradicts the latter study’s dichotomization approach.

A large two-sample Mendelian randomization-based study (28) showed a significant association between higher IL-6 signaling and suicidality. No statistically significant association was found between other depressive symptoms and higher CRP levels or IL-6 signaling. The study showed, however, that anhedonia, tiredness, feelings of inadequacy, and changes in appetite were significantly associated with higher BMI. (28) Depression symptoms were assessed using the Patient Health Questionnaire-9 (PHQ-9), and the study did not differentiate between cases with and without major depression, both eligible, as in our cohort. However, no age ranges were reported, and it remains unclear whether the results might be replicated in older populations. The study confirms our results regarding the lack of association between CRP levels and depression-specific features in the total study population. However, in our study, measures of the Abdominal circumference were more promising in estimating anhedonic features than BMI when we decreased the number of cases in groups after depression-specific stratification. Furthermore, the contradictory results regarding the association between IL-6 and anhedonia between our study and the latter pose questions about whether anhedonic features captured in our analysis might be a prior stage or proxy of higher risk of suicidal ideation. GDS does not directly explore the presence of suicidal ideas. However, one question (GDS 15: “Do you think it is wonderful to be alive now?”) might present some indirect annotation and was incorporated into the anhedonic features in our study. Furthermore, many questions referring to tiredness and feelings of inadequacy were classified into anhedonic and melancholic features in our study; both clusters were significantly associated with high IL-6.

Combining data from the UK-Biobank and NESDA showed that IL-6 but not CRP was associated with PHQ-9-related anhedonia. (29) Higher NESDA-measured IL-6 levels were additionally associated with higher odds of hypersomnia, fatigue, and decreased appetite. Higher CRP levels, however, were associated with increased odds of fatigue, sleep problems, depressed moods, and irritability after a meta-analytic pooling of estimates across the two cohorts. Despite maintaining their statistical significance, these latter associations were attenuated after adjusting for BMI. In the Mendelian Randomization analysis, genetically predicted higher CRP levels were associated with suicidal ideation and cognitive problems. The genetically predicted high IL-6 signaling was associated with fatigue and sleep alterations. (29)

A network analysis-based study on polygenic risk scores of immunometabolic biomarkers and depression symptoms showed an association between CRP and changes in appetite, fatigue, and anhedonia. (18) Contrary to previous findings, CRP and anhedonia exhibited a negative association. Furthermore, the study showed a significant association between TNF-alpha and fatigue and between BMI, anhedonia, and changes in appetite. (18) Despite the interesting findings, the study has several limitations. The combination of clinical and population-based cohorts, the use of different assessment tools for depression symptoms across different cohorts (Hamilton Rating Scale for Depression and PHQ-9), the lack of IL-6 and IL-10 specific assessments in one of the included cohorts (selection bias), and, as previously mentioned, the restrictiveness to participants with European ancestry present major limitations. The inclusion of younger age groups (mean ages: 47.8, 43.2, and 56.2 years) limits the generalizability of the results to older populations. (18)

Clustering patients based on CRP levels and “difficult-to-treat depression” criteria in a clinical study (37.9±13.3, n=263 with depression) showed higher inflammatory biomarkers (IL-6 and IL8), and higher BMI in those with CRP > 3mg/L. (22) The exclusion of cases with inflammatory disorders did not change the results. However, no significant difference in the Montgomery-Asberg Depression Rating Scale (MADRS) or suicide assessment scale (SUAS) scores was found between the groups with higher and lower CRP levels. (22)

### 4.2. Diabetes

The Irish Longitudinal Study on Aging-based analysis in a large cohort of middle-aged and older adults (≥ 49 years, n=3,895) showed a significant association between higher CRP levels and depression in T_2_DM participants but not in controls. (30) Furthermore, only in those with T_2_DM, CRP levels at baseline predicted higher depression risk after a 4-year follow-up. (30) In a large Japanese cross-sectional study on middle-aged and older adults with T_2_DM (66±11.4 years, n=3,573), higher CRP levels were associated with 169% higher odds of having depression only in those with a BMI ≥ 25 (recommended cutoff for obesity in the Japanese population). (31) In the same study, stratifying by HbA1c levels did not show significant results. (31) The depression diagnosis was based on nine items from the PHQ-9. The authors did not exclude a potential involvement of insulin resistance in the observed results, and acknowledged the limitation associated with the lack of this information. (31) A smaller study in middle-aged participants (48.1±12 years, n=219) with poorly controlled diabetes found a significant value of baseline CRP levels in predicting a lower improvement of depression symptoms after treatment in T_1_DM but not T_2_DM. (32) The lack of data on IL-6 levels in these studies limits the interpretation of these results, and the differences between populations and severity of diabetes might explain observed discrepancies.

### 4.3. Obesity

Our previous study has shown a strong association between depression, IL-6 levels, and obesity. IL-6 was a strong mediator in the association between depression and obesity. (15) The association between depression, BMI, and inflammation has largely been investigated, and studies have shown conflicting results. A 2009-2010 National Health and Nutrition and Survey (NHANES) based study showed that in 585 participants with depression (48.74±15.88 years), higher CRP levels (3 and 5 mg/L as cutoffs) were significantly associated with higher BMI and higher odds of metabolic syndrome (2.81 and 1.94, respectively). (33) Depression was assessed using PHQ-9. In smaller UK-based research in middle-aged adults (48.7±15.1 years, n=406), the associations between the polygenic risk score of depression and IL-10 levels, and the polygenic risk score of BMI and IL-6 and CRP levels did not survive the adjustment for multiple testing. (34)

Another NESDA-based study (40.96±12.25 years, n=1,077) showed a significant association between CRP and atypical, energy-related symptoms (AES= increase in appetite, weight gain, fatigue, hypersomnia, and leaden paralysis) in patients with current major depression. (35)

The study found a significant dose-related association between CRP, AES, and different immune-metabolic biomarkers (IL-6, TNF-alpha, Glycoprotein acetyls, BMI, Waist circumference, Triglycerides, Glucose, Leptin, and HDL cholesterol). (35) While this study adjusted for the use of antidepressant medication, which presents a strength in the analysis, CRP, on which the study was based, is a nonspecific biomarker of inflammation, as the contradictory results of previous studies have shown.

### 4.4. Strengths

This is the first study to explore the association between the immune-metabolic and depression-associated features specifically in middle-aged and older adults of multi-ethnic background. The large number of included participants and the high availability of inflammation biomarkers present a major strength of the current analysis. The studied biomarkers are clinically relevant and might be assessed in everyday practice, and the assessment of IL-5, IL-6, IL-10, and TNF-alpha is a further strength compared to studies that focused only on CRP. The use of the GDS score, a clinically relevant diagnostic tool with high sensitivity in older adults, is a further novelty compared to available data. The application of different methods to highlight the associations between the variables and the exploration and visualization of relationships within and between clusters is a novelty. The inclusion of a large sample of ethnic minorities, with higher risks of stress, depression, and cardiometabolic disorders, in addition to the community-based aspect of the source cohort, allows for more generalizability of the results in ethnically diverse settings of community-dwelling adults. Finally, the current findings were adjusted for multiple confounders not explored in previous publications, mainly the effect of anti-inflammatory, statin, antidepressant, and antidiabetic medications. The findings were also adjusted for the effect of cognitive decline since apathy and anhedonia might be independently reported in the course of dementia. (36) This reflects the robustness of the presented associations.

### 4.5. Limitations

The missing values in some immunological biomarkers present a major limitation. CRP levels were included for the comparability of our data with previous publications, but were not assessed in African-American participants. This presents a selection bias toward this variable, and the corresponding results might not apply to African-American populations. Besides CRP, the included number of participants remained significantly high compared to previous cohorts, and correlation analyses were performed pairwise to avoid loss of important information in the data. The community-based aspect of the cohort and the non-random sampling of study participants might present a selection bias by including socially active individuals. However, this recruitment strategy allowed reaching ethnic groups rarely covered by clinical studies and randomized trials. The focus of the current analysis was on community-dwelling middle-aged and older adults with a higher representation of ethnic minorities, and it does not reflect potential findings in more severely affected persons, generally captured in geriatric clinical studies. Furthermore, the median educational level was 14 years, which indicates high participation rates of well-educated individuals, and the study might not reflect findings in more deprived communities. Clinical diagnoses were based on self-disclosure, and no clinical records were revised. However, this might present a very low risk to the significance of our results, seeing that the main analyses and results were based on biological and neuropsychological assessments performed by certified study clinicians. The cross-sectional design is a further limitation, since we cannot draw a causal conclusion from the current results. Subsequent assessments were made at 24–30 months, and many participants were lost to follow-up owing to the COVID pandemic and the non-clinical aspect of the study. The current study focused more on understanding clusters and drawing concomitant networks between them rather than exploring causal associations. Longitudinal frameworks are exposed to both within and between individual variability in the inflammatory and metabolic biomarkers and do not present the best design to answer the current study question, based on associations and not causation. Finally, no information is available on prior variations of the variables of interest.

## 5. Conclusions

This correlation network-based analysis of a large multiethnic middle-aged and older population is in favor of an overlap between immunological, metabolic, and specific depression-related features. These features are neither specific nor restricted to depression, as the correlations between IL-6, Abdominal circumference, and anhedonic features were maintained in the subgroup without current depression. This study is a first step toward clustering different subtypes of immunometabolic depression-associated symptoms for a better risk stratification and to direct personalized preventive and therapeutic strategies.

## Declarations

### Conflicts of interest

The authors have no conflict of interest, neither financial nor non-financial.

Ethical approval:

All procedures contributing to this work comply with the ethical standards of the relevant national and institutional committees on human experimentation and with the Helsinki Declaration of 1975, as revised in 2013. Ethical approval was obtained from the local institutional review board. Participants gave written informed consent. The current research is based on a secondary analysis of anonymized data and was performed in compliance with the data use agreement (DUA with Hallab).

### Authorization for publication

The principal investigator and data administrator of the HABS-HD study reviewed the manuscript for its compliance with DUA and authorized the submission and publication of the current version.

### Authorship

AH has full access to all of the data and takes responsibility for the integrity of the data and the accuracy of the analysis, visualization, drafting, and editing of the manuscript.

### Data availability

Data can be acquired by qualified researchers after an official request.

